# Longitudinal analysis of SARS-CoV-2 seroprevalence using multiple serology platforms

**DOI:** 10.1101/2021.02.24.21252340

**Authors:** Juan Manuel Carreño, Damodara Rao Mendu, Viviana Simon, Masood A Shariff, Gagandeep Singh, Vidya Menon, Florian Krammer

**Affiliations:** Dept. of Microbiology, Icahn School of Medicine at Mount Sinai, New York, NY, USA; Clinical Microbiology Laboratory, Department of Pathology, Icahn School of Medicine at Mount Sinai, New York, USA; Department of Internal Medicine, NYC Health + Hospitals/Lincoln, The Bronx, NY, USA; Division of Infectious Diseases, Department of Medicine, Icahn School of Medicine at Mount Sinai, New York, NY; The Global Health and Emerging Pathogens Institute, Icahn School of Medicine at Mount Sinai, New York, NY

## Abstract

Serological tests are important tools helping to determine previous infection with severe acute respiratory disease coronavirus 2 (SARS-CoV-2) and to monitor immune responses. The current tests are based on spike (S), the receptor binding domain (RBD), or the nucleoprotein (NP) as substrate. Here, we used samples from a high seroprevalence cohort of health care workers (HCWs) to perform a longitudinal analysis of the antibody responses using three distinct serological assays. 501 serum samples were tested using: a) a research-grade RBD and spike based tandem enzyme-linked immunosorbent assay (MS–RBD ELISA, MS-spike ELISA), b) a commercial RBD and spike based tandem ELISA (Kantaro-RBD, -spike), and c) a commercial NP-based chemiluminescent microparticle immunoassay (CMIA, Abbott Architect). Seroprevalence ranged around 28% during the early stage of the pandemic (a: 28.4% positives; b: 28.1%; c: 27.3%). Good correlation was observed between the MS and Kantaro RBD ELISAs and between the MS and Kantaro spike ELISAs. By contrast, modest correlations were observed between the Abbott Architect and both RBD and spike-based assays. A proportion of HCWs (n=178) were sampled again 3-5 months after the first time point. Although antibody levels declined in most of the positive individuals, the overall seroprevalence measured by RBD-spike based assays remained unchanged. However the seroprevalence of NP-reactive antibodies significantly declined. Lastly, we tested six samples of individuals who received two doses of SARS-CoV-2 mRNA vaccine and found that seroconversion was detected by the RBD-spike assays but – of course as expected - not the NP based assay. In summary, our results consolidate the strength of different serological assays to assess the magnitude and duration of antibodies to SARS-CoV-2.

## Introduction

In the advent of the current pandemic caused by the severe acute respiratory syndrome coronavirus 2 (SARS-CoV-2), pandemic, methods to detect the prevalence of recent and past infections are key to determine public health and social countermeasures. Nucleic acid amplification tests (NAAT) provide an accurate estimation of acute infections (1, 2), but they fail to inform about past infections. Serological tests that detect antibodies directed against structural targets of the virus, not only are useful to estimate the overall viral seroprevalence and rates of infection in the population (3-5), but also help to assess responses to vaccination (6), to determine correlates of protection (7, 8), and to test and standardize therapeutic approaches such as monoclonal antibody and plasma transfer therapies (9). Moreover, estimation of viral seroprevalence and quantification of antibody levels adds to our understanding of the immune response and protection at the individual and population levels (10).

Currently, serological assays to detect antibodies against SARS-CoV-2 are based on recombinant versions of the spike (S), the receptor binding domain (RBD) of S, or the nucleoprotein (NP) as substrate (11). A variety of research grade and commercial S-based and NP-based assays are now available, but antibodies to these two targets have different characteristics. Antibodies directed against the viral spike are retained for several months after infection (12-17) and correlate with virus neutralization and protection against re-infection (6, 13, 18-21). Moreover, vaccination relies uniquely on the viral spike, evidencing the importance of detecting antibodies against this target with high levels of sensitivity and specificity (6).

Several studies evaluate the sensitivity and specificity of individual assays, either S- or NP-based, however longitudinal side-by-side comparisons of different serological platforms are scarce. Here, we employed samples from a high-risk cohort of health care workers (HCWs) using three different serological assays. In addition, SARS-CoV-2 post-vaccination samples were included in the analysis. We compared a research grade RBD and spike based tandem enzyme-linked immunosorbent assay (ELISA) developed at Mount Sinai (MS ELISA, research grad version), the Seroklir commercial RBD-spike based ELISA from Kantaro Biosciences, and the commercial NP-based chemiluminescent microparticle immunoassay (CMIA) Abbott Architect.

## Methods

### Research grade ELISAs

Detection of receptor binding domain (RBD) and full-length spike (S) antibodies in plasma was performed with a research-grade two-step ELISA developed at Mount Sinai closely resembling an assay used in Mount Sinai’s CLIA-certified Clinical Pathology Laboratory, which received FDA Emergency Use Authorization in April 2020 (22, 23). The research grade assay has 95% sensitivity and 100% specificity (3). Before performing the ELISA, samples were heat-inactivated for 1h at 56°C. Briefly, for RBD screening, 96-well plates (Thermo Fisher) were coated with 50ul/well of phosphate-buffered saline (PBS; Gibco) containing 2μg/ml of recombinant RBD protein and incubated overnight at 4 °C. Plates were washed three times with PBS containing 0.1% Tween-20 (PBS-T; Fisher Scientific) using an automated plate washer (BioTek). For blocking, 200μl/well of PBS-T containing 3% (w/v) of milk powder (American Bio) were added and plates were incubated for 1 h at room temperature. Plasma samples were diluted (1:50) in PBS-T containing 1% milk powder. Blocking solution was removed and dilutions of samples were added. After a 2-hour incubation, plates were washed three times with PBS-T and 50μl/well of anti-human IgG (Fab-specific) horseradish peroxidase antibody (Sigma, A0293) diluted 1:3,000 in PBS-T 1% milk powder were added. Plates were incubated for 1 h at room temperature, followed by three times washing with PBS-T and addition of developing solution (100μl/well) sigmafast *o*-phenylenediamine dihydrochloride (Sigma). The reaction was led to proceed for 10 min, and stopped using 50μl/well of 3-molar hydrochloric acid (Thermo Fisher). Optical density was measured at 490 nm using an automated plate reader (BioTek). Samples with an OD_490nm_ above 0.15 (cut-off value) were considered as presumptive positives and were further tested in the confirmatory ELISA using the full-length recombinant spike protein.

Briefly, to perform the confirmatory ELISAs, plates were coated and blocked as described above, but using full-length spike protein for coating. Presumptive positive plasma samples were serially diluted (1:3) in 1%-milk prepared in PBS-T, starting at an initial dilution of 1:80. Serial dilutions (100μl/well) were added to the plates, followed by 2-hour incubation at room temperature. The remaining steps were performed as described above. Data was analyzed using GraphPad Prism 7. Samples with an OD_490nm_ above 0.15 (cut-off value) at a 1:80 plasma dilution were considered positive. Samples with an OD_490nm_ above 0.15 at the last dilution were further diluted (1:2160 initially) and re-tested. Only samples positive in both steps of the assay were considered positive.

### Kantaro ELISAs

ELISAs to detect antibodies in plasma against the receptor binding domain (RBD) and the full-length spike (S) based on the commercial Kantaro Quantitative SARS-CoV-2 IgG Antibody Kit (COVID-SeroKlir, Kantaro Biosciences) were used. The assay was performed according to manufacturer’s instructions except for additional serum dilution steps in highly reactive individuals. All reagents and microplates were included with the commercial kit. Briefly, for qualitative RBD ELISAs, samples were diluted in sample buffer (1:100) using 96-well microtitre plates, and 100μl/well of pre-diluted samples were transferred to the RBD pre-coated microplates. Positive and negative controls were added to every plate. Samples were incubated for 2 hours at room temperature, followed by removal of plasma dilutions and washing three times with wash buffer. RBD conjugate was diluted in conjugate buffer and 100μl/well were added to the plates. After 1h incubation, conjugate was removed and plates were washed three times with wash buffer. Substrate solution was added (100μl/well) and after 20min incubation, 100μl/well of stop solution were added. Samples were read at OD_450nm_ and at OD_570nm_ for wavelength correction. The cutoff index (CI) was calculated by dividing the corrected OD of the clinical sample/corrected OD of RBD positive control. Samples with a CI above 0.7 were considered as presumptive positives and were further tested in the confirmatory quantitative ELISA based on the full-length recombinant spike protein.

For quantitative spike ELISAs, presumptive positive plasma samples were diluted (1:200) in sample buffer. Dilutions were added in duplicate to the pre-coated microplates. Low, medium and high controls, as well as spike calibrators used to generate a standard curve, were added to every microplates. After 2h incubation at room temperature, the remaining steps were performed as described above. Data was analyzed using GraphPad Prism 7. Concentration of spike-reactive antibodies was calculated using a four parameter logistic (4-PL) curve fit. Samples exceeding the range of the standard curve were further diluted (1:5400) and re-tested. Only samples positive in both steps of the assay were considered positive.

### Abbott Architect CMIA

The Architect test (Abbott Laboratories) consists of an automated, two-step, qualitative CMIA for qualitatively detecting IgG against the nucleoprotein (N) antigen from SARS-CoV-2. This test has a reported sensitivity of 100% (CI 95.8–100%) and specificity of 99.6 (CI 99–99.9%) 14 days after symptom onset. The assay was performed according to manufacturer’s instructions. All reagents were included with the kit. Briefly, sample, SARS-CoV-2 antigen coated paramagnetic microparticles, and assay diluent were combined and incubated. The IgG antibodies to SARS-CoV-2 present in the sample bind to the SARS-CoV-2 antigen coated microparticles. The mixture is washed. Anti-human IgG acridinium-labeled conjugate is added to create a reaction mixture and incubated. Following a wash cycle, Pre-Trigger and Trigger Solutions are added. The resulting chemiluminescent reaction is measured as a relative light unit (RLU). There is a direct relationship between the amount of IgG antibodies to SARS-CoV-2 in the sample and the RLU detected by the system optics. This relationship is reflected in the calculated Index (S/C). The presence or absence of IgG antibodies to SARS-CoV-2 in the sample is determined by comparing the chemiluminescent RLU in the reaction to the calibrator RLU.

### Study participants and human samples

The samples used for the longitudinal study, were part of a cross sectional cohort of healthcare workers (HCWs) of the New York City Public Hospital in the South Bronx. This study was approved by the Institutional Review Board (IRB#20-009). Samples were collected in two phases: Phase 1 samples were obtained in May 2020 and Phase 2 samples were collected from August to October 2020. Informed consent was obtained prior to Phase 1 sample collection.

Samples from study participants receiving the Pfizer mRNA vaccine were obtained from IRB approved longitudinal observation studies (IRB-16-00791; IRB-20-03374) conducted by the Personalized Virology Initiative (PVI) at the Icahn School of Medicine at Mount Sinai. All participants signed informed consents prior to data and sample collection. All serum samples were coded upon collection and analyzed in a blinded manner in the Krammer laboratory.

### Statistical analysis

Correlations of antibody levels in the different assays were calculated using a standard Pearson’s correlation. Coefficients of correlation (r) are presented. Paired t-test for comparison of phase 1 and phase 2 antibody levels was used. All adjusted P values of <0.05 were considered statistically significant with a confidence interval of 95%. Statistical analyses were performed using Prism 7 (GraphPad, USA).

## Results

### Longitudinal comparison of SARS-CoV-2 seroprevalence using RBD/spike and NP based assays

Seroprevalence of SARS-CoV-2 across different regions of the world has been described using multiple serological assays based either on the spike protein (S), its receptor-binding domain (RBD), or the nucleoprotein (NP). Here we compared side-by-side a research grade MS ELISA based on RBD and spike, an RBD/spike based SeroKlir assay from Kantaro Biosciences and the NP based Abbott Architect test. We used a set of 501 samples from frontline healthcare workers (HCW) collected after the first pandemic wave in the New York City area (May 2020). Seroprevalence in this set of samples using a research grade ELISA from Mount Sinai was 28.4%, (142/501), 28.1%, using the SeroKlir test from Kantaro Biosciences (141/501) and 27.3% using the Abbott Architect test (137/501) (**Fig. 1A**). A subset of the initial participants (n=178) attended provided a second serum sample at a follow up visit in August-October 2020 allowing assessment of seroprevalence at two different time points. Of note, the seroprevalence in the smaller subset of participants was higher compared to the initial cohort (N=501). This is likely due to higher compliance of participants that knew their sero-status in the first phase, since they were informed about their antibody levels. Overall, the seroprevalence measured by the Mount Sinai and the Kantaro ELISAs did not vary significantly between the two time points (**Fig. 1B-1C**), but the seroprevalence of NP reactive antibodies measured by the Abbott Architect test declined (**Fig. 1D**).

**Figure 1.**
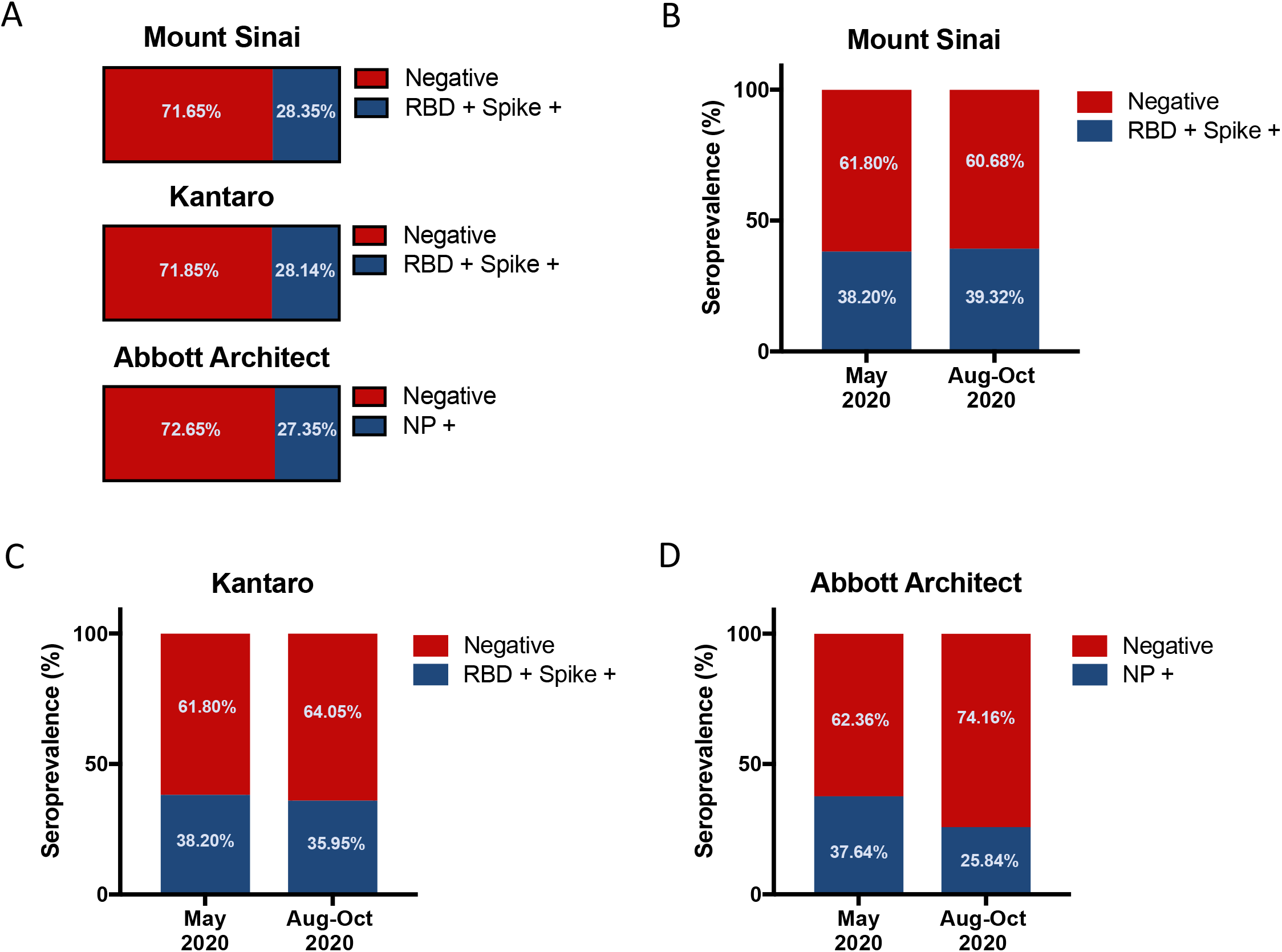
Seroprevalence against SARS-CoV-2 in healthcare workers using different serology platforms. Serum samples from frontline healthcare workers were assessed for antibodies against RBD-Spike using a research grade ELISA from Mount Sinai, against RBD-Spike using a commercial ELISA from Kantaro Biosciences, or against NP using Abbott-Architect CMIA. Initially, samples analyzed in the three assays consisted of specimens obtained early during the pandemic on May 2020 (n=501) (**A**). Seroprevalence in a subset of subjects (n=178) who attended a follow up visit on August-October 2020 was determined using the three different serological assays and a comparison of the two time points is shown (**B-C**).

We further compared the antibody levels in samples obtained during the first phase (May 2020) and the second phase (Aug-Oct, 2020) in the subset of 178 subjects (the distribution of antibody levels is shown in **Sup. Fig. 1** and concordance analyses among the different assays are shown in **Sup. Fig. 2**). As expected, antibody levels in the second phase declined in the majority of participants in a manner that was consistent in the three different assays (Figs. **2A-2C**). A sharper decline of NP reactive antibody levels as measured by the Abbott Architect test (Fig. **2C**). Moreover, the percentage of subjects that were seropositive initially and whose antibodies became undetectable in the second phase did not vary significantly in the Mount Sinai and Kantaro ELISAs (**Figs. 2A-2B**) but 30% of the samples that were positive initially in the Abbott Architect test became negative in the second phase (**Fig. 2B**).

**Figure 2.**
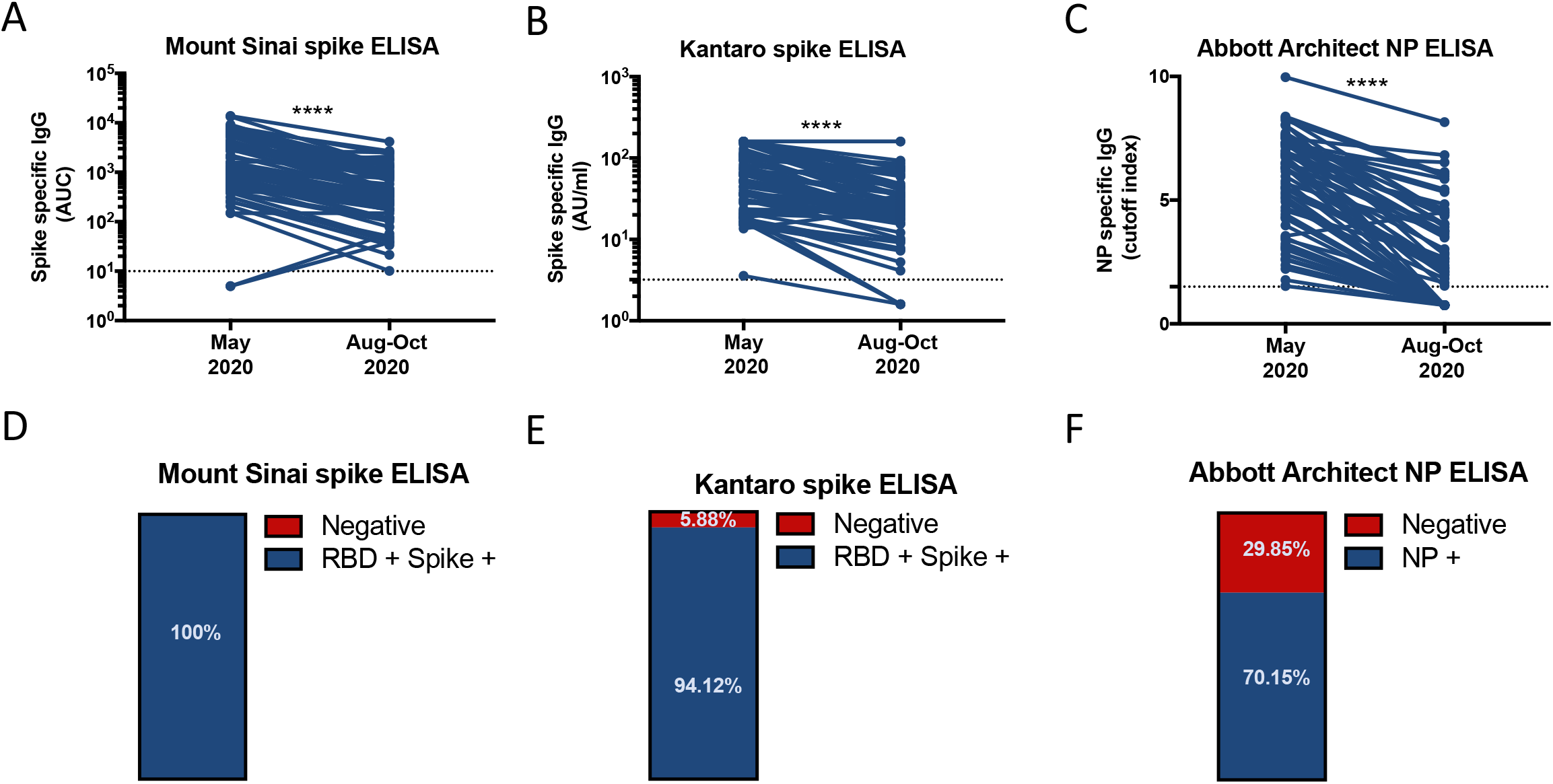
Longitudinal analysis of antibody levels against RBD-Spike or NP in seropositive subjects. Serum samples from frontline healthcare workers were obtained on May 2020 or August-October 2020. Antibodies against RBD-Spike were measured using a research grade ELISA from Mount Sinai (**A**); against RBD-Spike using a commercial ELISA from Kantaro Biosciences (**B**); or against NP using the Abbott-Architect CMIA (**C**). Antibody levels in specimens obtained early during the pandemic on May 2020 or in a follow up visit on August-October 2020 are shown. Samples with a value above or below the cutoff of the corresponding assay (doted line) are shown. **** P<0.0001. The percentage of seropositive samples that turned negative (red) or that remained positive (blue) as measured in each of the corresponding assays is shown in **D-F**.

### Correlation of antibody levels among the different assays

The antibody response against different antigenic targets of a particular virus exhibits a high degree of complexity. The magnitude and kinetics of the antibody response against RBD/spike and the nucleoprotein are not fully understood. To analyze the consistency between the two RBD/spike based assays and to study the relationship of RBD/spike reactive antibodies versus NP reactive antibodies, we performed correlation analyses among the three different assays. Using the positive samples from the first (**Figs. 3A-C**) and second (**Figs. 3D-2F**) phases, we detected a good correlation of RBD reactive antibodies (optical density (OD) measured at one dilution) measured by the Mount Sinai ELISA versus the Kantaro ELISA either in phase 1 (r= 0.9169; P two-tailed= <0.0001, **Fig. 3A**) or phase 2 (r= 0.9075; P two-tailed= <0.0001, **Fig. 3D**). However, the correlation of RBD reactive antibodies measured in the Mount Sinai or Kantaro assays versus the NP reactive antibodies measured in the Abbott Architect test, either in phase 1 (**Figs. 3B-3C**) or phase 2 (**Figs. 3E-3F**) samples, was modest to low.

**Figure 3.**
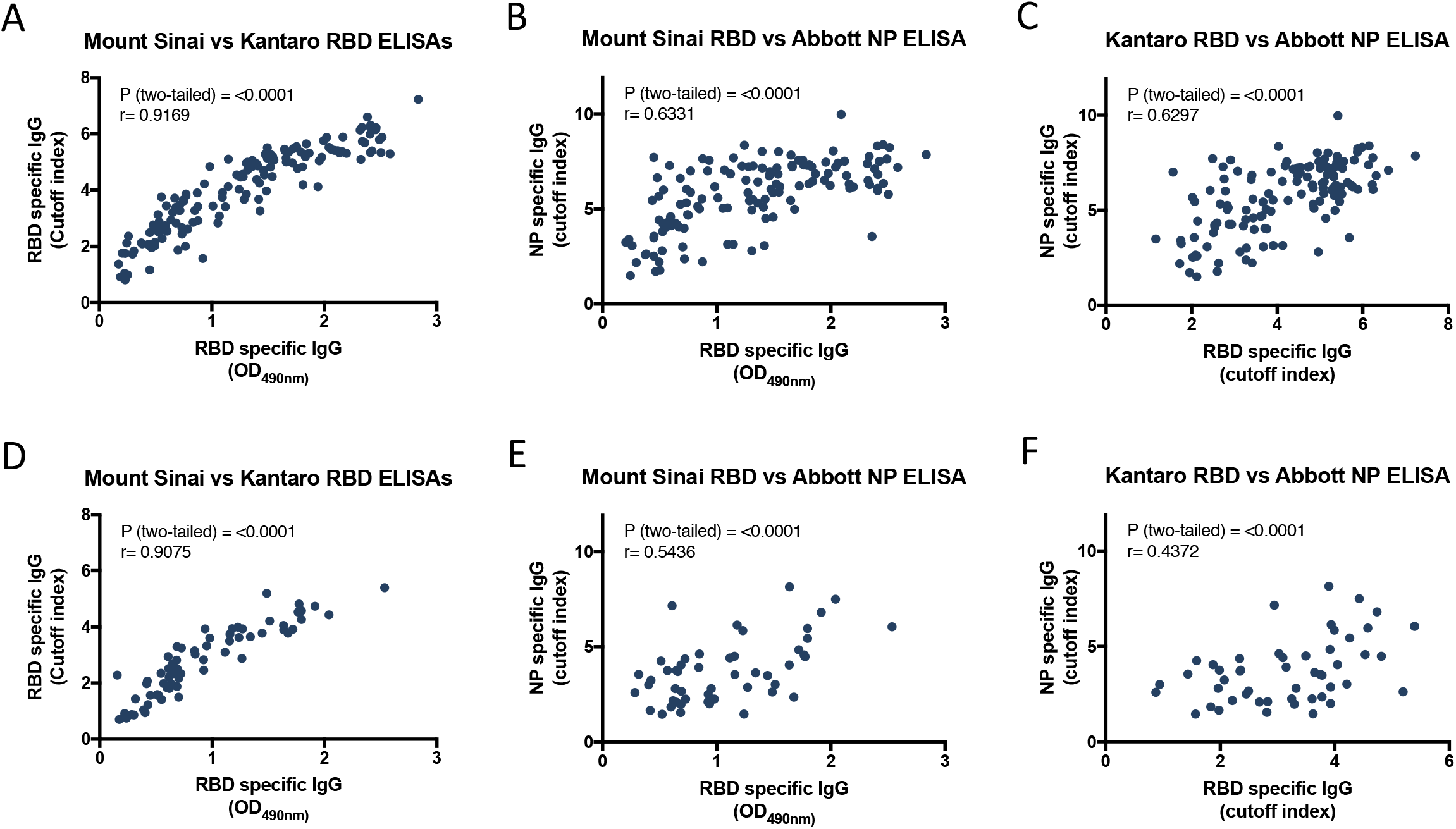
Correlation analysis of antibody levels (RBD-reactive) among the different serology assays. Serum samples were assessed for antibodies against RBD-Spike using a research grade ELISA from Mount Sinai, against RBD-Spike using a commercial ELISA from Kantaro Biosciences or against NP using the Abbott-Architect CMIA. Correlation of antibody levels (RBD reactive) among the different assays using serum samples obtained on May 2020 (first time point) is shown (**A-C**). Correlation of antibody levels (RBD reactive) among the different assays using serum samples obtained on August-October 2020 (second time point) is shown (**D-F**). Correlation analysis between Mount Sinai RBD and Kantaro RBD ELISAs (**A, D**); between Mount Sinai RBD and Abbott Architect NP ELISAs (**B, E**); and between Kantaro RBD ELISAs and Abbott Architect NP CMIAs (**C, F**) are shown. Pearson correlation was used. Significance and coefficient of determination are shown.

We performed next the same type of analyses but with quantitative spike reactive antibody levels instead of RBD reactive antibody OD values. Again, we found a good correlation between the Mount Sinai ELISA vs the Kantaro ELISA either in phase 1 (r= 0.6860; P two-tailed= <0.0001, **Fig. 4A**) or phase 2 (r= 0.9135; P two-tailed= <0.0001, **Fig. 4D**) and a weak correlation between spike reactive antibodies measured in the Mount Sinai or Kantaro assays versus the NP reactive antibodies measured in the Abbott Architect test (**Figs. 4B-4C:** phase 1; **Figs. 4E-4F:** phase 2). For both RBD and spike, some of the subjects exhibited very high levels of RBD reactive antibodies and low levels of NP reactive antibodies and *vice versa*. Overall, these findings indicate that the magnitude of RBD/spike and NP antibody responses differs considerably highlighting the need for further studies using samples from well-described longitudinal cohort studies.

**Figure 4.**
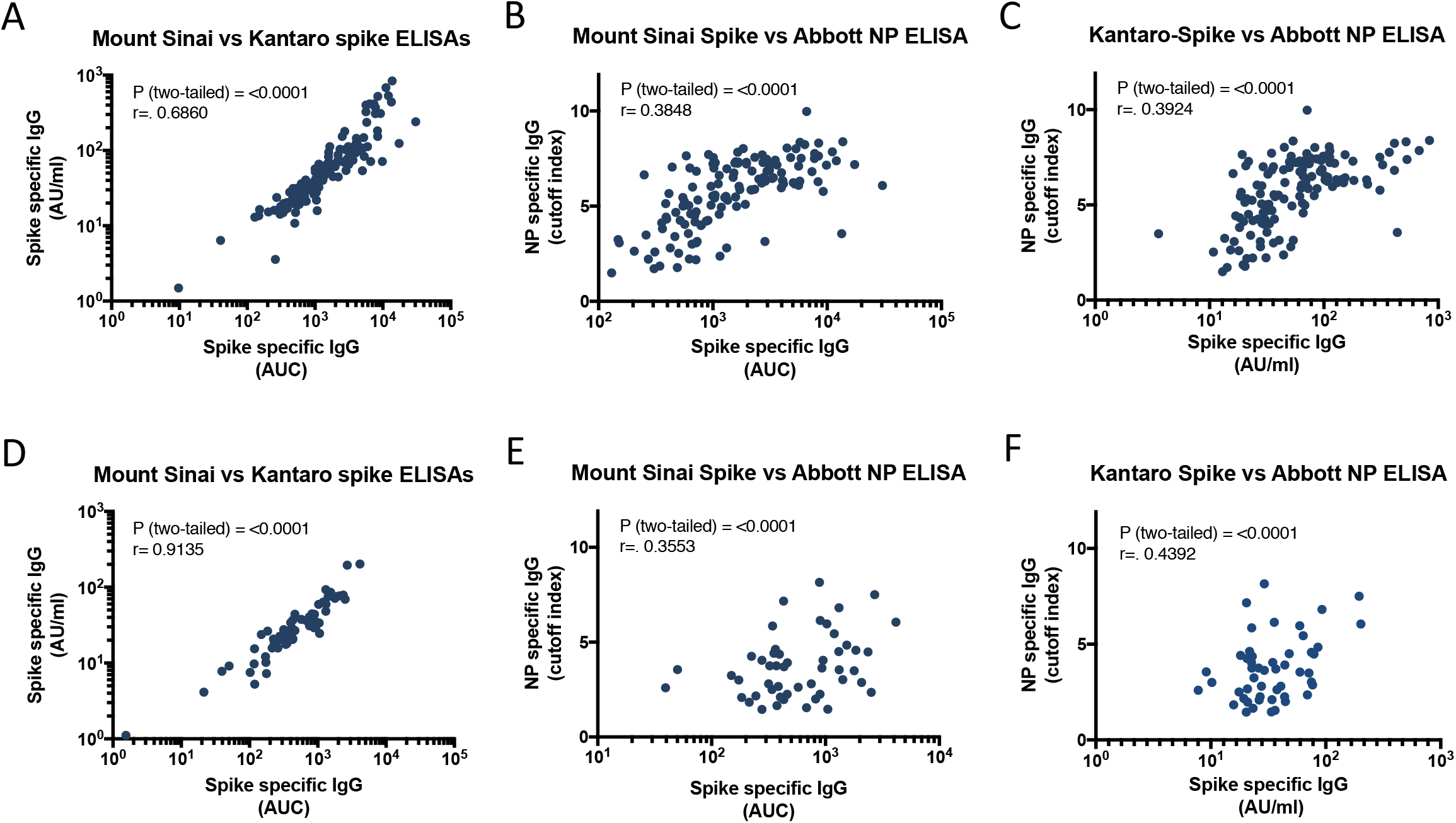
Correlation analysis of antibody levels (spike-reactive) among the different serology assays. Serum samples were assessed for antibodies against RBD-Spike using a research grade ELISA from Mount Sinai, against RBD-Spike using a commercial ELISA from Kantaro Biosciences or against NP using the Abbott-Architect CMIA. Correlation of antibody levels (spike reactive) among the different assays using serum samples obtained on May 2020 (first time point) is shown (**A-C**). Correlation of antibody levels (spike reactive) among the different assays using serum samples obtained on August-October 2020 (second time point) is shown (**D-F**). Correlation analysis between Mount Sinai RBD and Kantaro RBD ELISAs (**A, D**); between Mount Sinai RBD and Abbott Architect NP CMIAs (**B, E**); and between Kantaro RBD ELISAs and Abbott Architect NP CMIAs (**C, F**) are shown. Pearson correlation was used. Significance and coefficient of determination are shown.

### Detection of vaccine induced antibodies in both assays

To date only limited data is available about how commercial antibody assays respond to antibodies developed in response to vaccination. In order to determine how the three assays perform against vaccine-induced antibodies, we measured reactivity in serum of individuals who had received two doses of SARS-CoV-2 mRNA vaccines. The expectation was, that the RBD/spike based assays would detect a signal while the NP based assay would not. Indeed, we measured high titers using the Spike based assay platforms (the Mount Sinai and Kantaro assays) but the samples produced no signal in the NP-based assay (**Fig. 5A-5C**). Of note, the spike titers measured in the Mount Sinai and Kantaro assay correlated very well (**Fig. 5D**).

**Figure 5.**
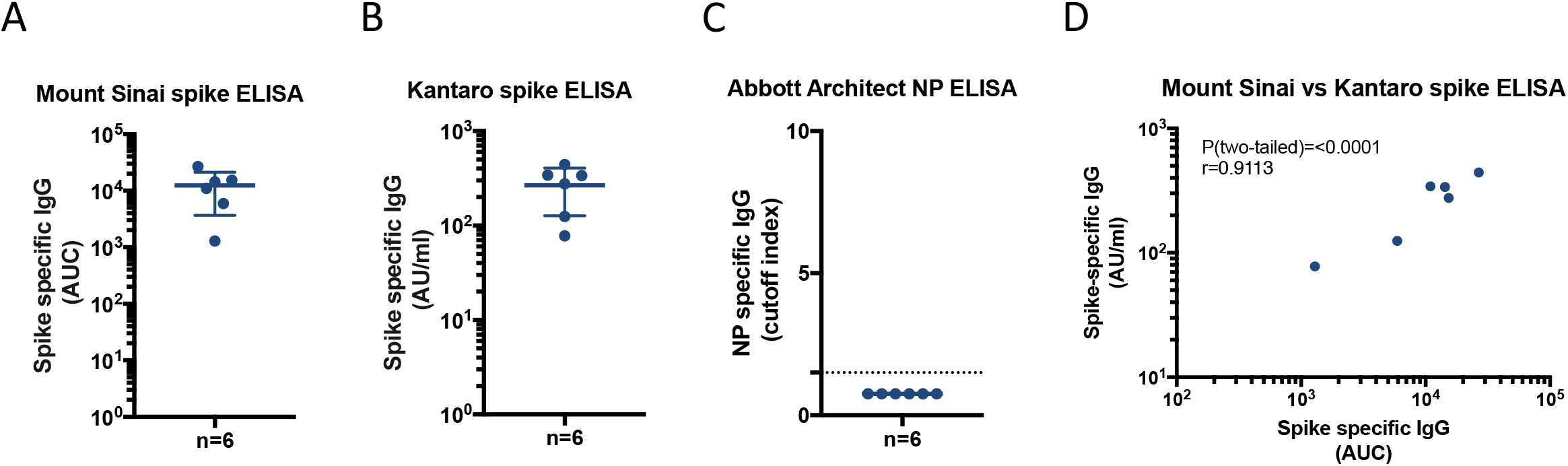
Post SARS-CoV-2 mRNA vaccination serum titers as measured in three different assays. Serum samples of individuals who had received two doses of SARS-CoV-2 mRNA vaccines were assessed for antibodies against RBD-spike using a research grade ELISA from Mount Sinai (A), against RBD-spike using a commercial ELISA from Kantaro Biosciences (B) or against NP using the Abbott-Architect CMIA (C). Correlation of antibody levels (spike reactive) between the Mount Sinai ELISA and the commercial ELISA from Kantaro Biosciences assays using the same serum samples (**D**).

## Discussion

While antibody responses to acute SARS-CoV-2 infection are relatively well understood, less data is available regarding antibody kinetics over longer time frames against different viral antigens. We determined seroprevalence and antibody titers in SARS-CoV-2 infected individuals at two time points (1-2 months and 3-4 months post infection) using three different assays. One assay, the Mount Sinai ELISA, is a laboratory-developed assay that uses an initial ELISA at a single serum dilution against the RBD followed by a confirmation and titration against the full-length spike protein. The second assay tested, the Kantaro SeroKlir assay, is based on the same principle, but commercially available. The third assay, the Abbott Architect, targets the NP and is a CMIA.

There was high concordance among the three assays with respect to seroprevalence during phase 1. However, the titers only correlated well for the two spike-based assays. During phase 2, the two spike-based assays identified all (Mount Sinai Research grade) or the vast majority (Kantaro) of previously seropositive individuals as seropositive, while the NP based assay (Abbott) failed to detect a signal above the cut-off in approximately 30% of previously positive individuals. These findings mirror similar results recently published by Grandjean and colleagues, suggesting that the NP antibody response is short-lived (24). However, it could also be a reflection of a high cut-off required to ensure high specificity for SARS-CoV-2 in the NP-based assay. Importantly, and as expected since no NP is included in the FDA EUA approved vaccines used in the US, only the spike-based assays were able to detect antibodies induced by SARS-CoV-2 mRNA vaccines (6). Our data highlight the need to understand assay performance before a specific assay is used to study specific aspects of SARS-CoV-2 immunity. All three assays are very valuable to assess seroconversion shortly after infection, but only the two spike-based assays were reliable months after recovery. Similarly, only spike-based assays are fit for measuring vaccine-induced antibodies, e.g. to determine if vaccination triggered immune responses.

## Data Availability

Data is available from the corresponding authors upon reasonable request.

## Acknowledgements

We thank the study participants for their continued generosity and willingness to support our longitudinal studies and our colleagues Adolfo Firpo and Carlos Cordon-Cardo for supporting our collaborative serology work. We also acknowledge the continued efforts of the Personalized Virology Initiative (in alphabetical order: Bulbul Ahmed, Hala Alshammary, Angela Amoako, Mahmoud Awawda, Katherine Beach, Carolina Bermúdez-González, Rachel Chernet, Lily Eaker, Shelcie Fabre, Emily. D. Ferreri, Daniel Floda, Charles Gleason, Dr. Giulio Kleiner, Dr. Denise Jurczyszak, Julia Matthews, Wanni Mendez, Dr. Lubbertus CF Mulder, Jose Polanco, Kayla Russo, Ashley Salimbangon, Dr. Miti Saksena, A. Shin, Levy Sominsky, Komal Srivastava, Sayahi Suthakaran). We thank research associates for their assistance with sample processing at Lincoln Hospital: Dominika Bielak, Ajmal Khan and Syed Hamad Ali Shah. Work in the Krammer and the Simon laboratories is partially funded by the NIAID Collaborative Influenza Vaccine Innovation Centers (CIVIC) contract 75N93019C00051, NIAID Center of Excellence for Influenza Research and Surveillance (CEIRS, contract # HHSN272201400008C), by the generous support of the JPB Foundation and the Open Philanthropy Project (research grant 2020-215611 (5384); and by anonymous donors. This effort was also supported by the Serological Sciences Network (SeroNet) in part with Federal funds from the National Cancer Institute, National Institutes of Health, under Contract No. 75N91019D00024, Task Order No. 75N91020F00003. The content of this publication does not necessarily reflect the views or policies of the Department of Health and Human Services, nor does mention of trade names, commercial products or organizations imply endorsement by the U.S. Government.

## Figure legends

**Supplementary Figure 1.**
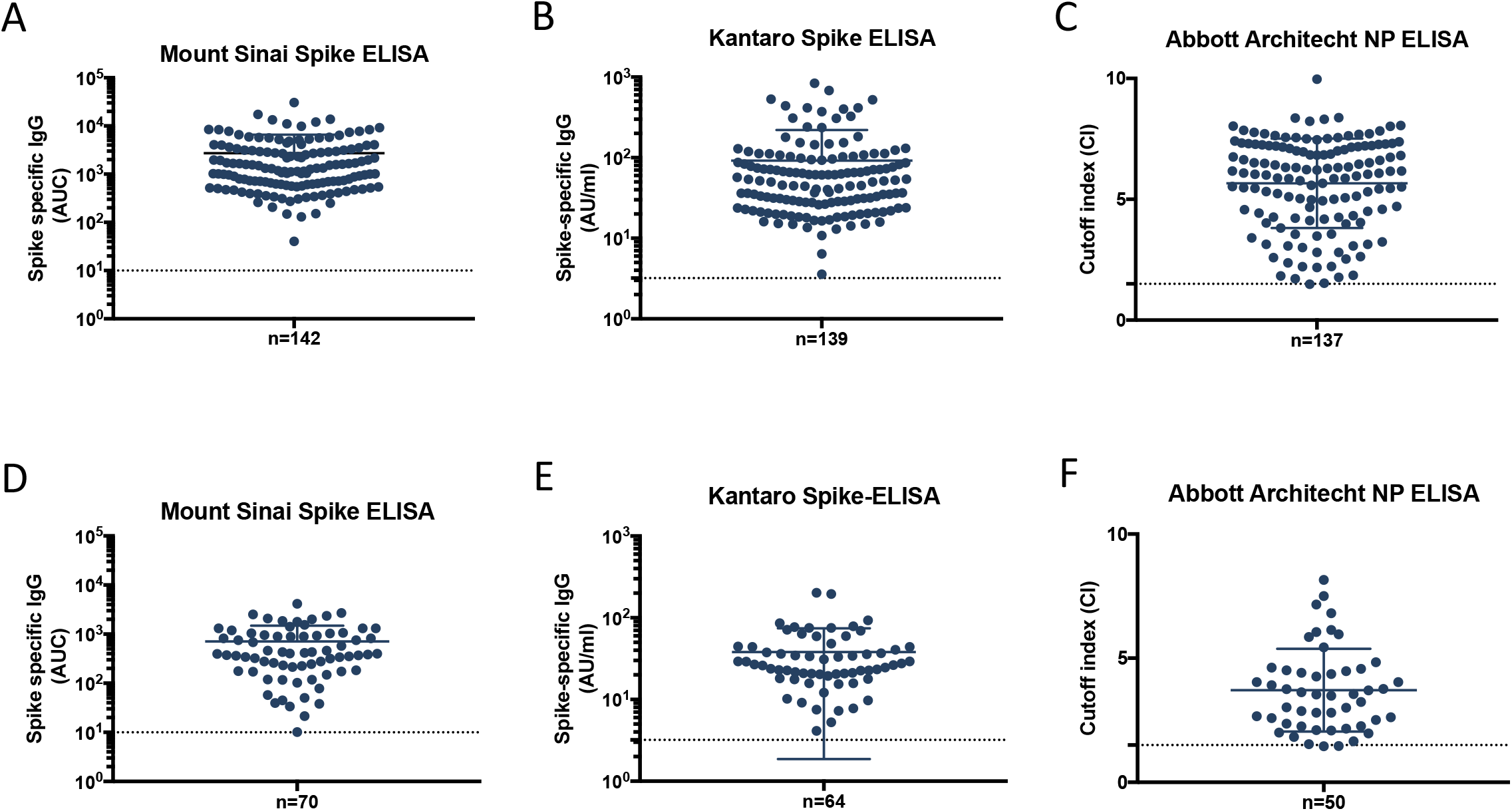
Distribution of antibody levels (spike-reactive) in the different serology platforms. Serum samples were assessed for antibodies against RBD-Spike using a research grade ELISA from Mount Sinai (A, D), against RBD-Spike using a commercial ELISA from Kantaro Biosciences (B, E), or against NP using the Abbott-Architect ELISA (C, F). Distribution of antibody levels (spike-reactive) early during the pandemic on May 2020 (**A-C**) or in August-October 2020 (**D-E**) are shown.

**Supplementary Figure 2.**
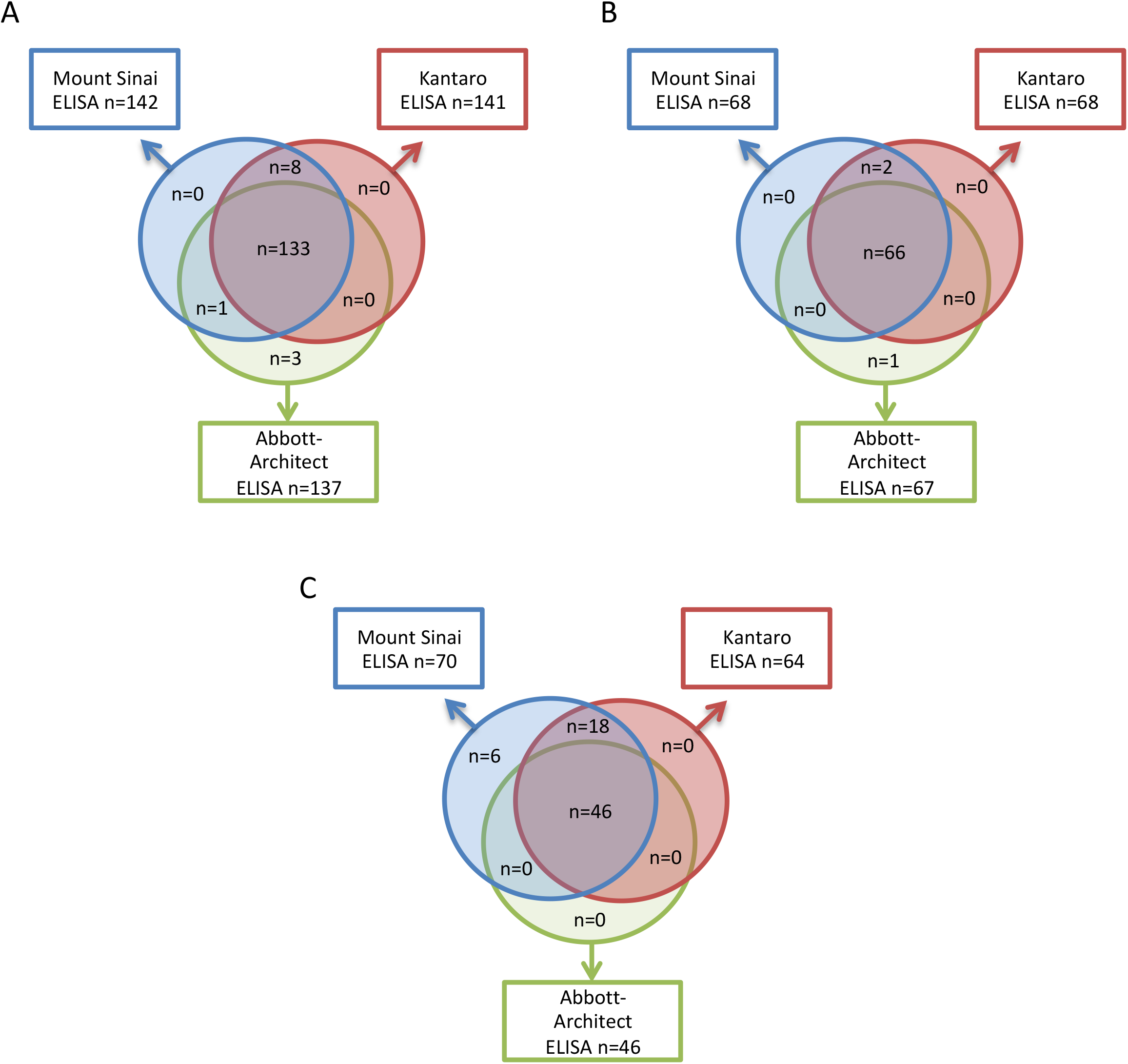
Concordance analysis of positive samples among the different serology assays. Venn diagrams depicting the number of seropositive samples measured by a RBD-Spike research grade ELISA from Mount Sinai (blue); a RBD-Spike commercial ELISA from Kantaro Biosciences (red), or an Abbott-Architect NP ELISA (green). Concordance of positivity among the three different assays using samples obtained early during the pandemic on May 2020 is shown (**A**). Seroprevalence in a subset of subjects (n=178) who attended a follow up visit on August-October 2020 was determined using the three different serological assays. Concordance of positivity in that subset of subjects on May 2020 (**B**) or August-October 2020 (**C**) is shown.

